# Co-designing CKD Talk Topics, a structured agenda-setting intervention for people with chronic kidney disease: “Keep It Simple, Stupid”

**DOI:** 10.64898/2026.10.28.26366231

**Authors:** Catherine H. Saunders, Anne Dade, Steve L. Bernstein, Clay A. Block, Hannah Crowe-Cumella, Glyn Elwyn, Lorraine Gerraughty, Gavin Henning, Vanessa Junkins, Ishaan Kumar, Annika Milliman, Joseph P. Nano, Geetha Ramkumar, Niveditta Ramkumar, Gabriela Sanders, Ailyn Sierpe, Quenton Turner-Gee, Anji Zhu, Ann O’Hare, JoAnna K. Leyenaar

## Abstract

**Rationale & Objective:** People with advanced chronic kidney disease (CKD) often face barriers to communication and self-advocacy, which may be mitigated by the practice of collaborative agenda-setting. We developed Chronic Kidney Disease Talk Topics (CKD Talk Topics), a structured agenda-setting intervention designed to help people with CKD and their care partners identify and communicate their discussion priorities during nephrology visits.

**Study Design:** Following a literature review, we used a participatory design approach involving semi-structured interviews with patients (with care partners who attended), and clinicians to develop CKD Talk Topics

**Setting & Participants:** Interviews occurred in a nephrology clinic in rural northern New England with patients that had advanced CKD (stages 4 and 5), some attending care partners, and clinicians.

**Analytical Approach:** We used Framework Analysis to summarize suggested content, format, and structure, and participatory Practical Thematic Analysis to explore communication needs and expectations.

**Results:** We interviewed 12 patients (with 4 accompanying care partners), and 8 clinicians. Overall, participants viewed CKD Talk Topics as a helpful tool. Patients emphasized simplicity, larger visual elements, CKD-specific content, and flexibility in usage. Clinicians noted the intervention’s potential for patient-centered, efficient visits. The final tool features six plain-language topics: “Your Body,” “Your Feelings,” “What Matters to You,” “Taking Care of Yourself,” “What Happens Next,” and “Everyday Needs.”

**Limitations:** Our findings may reflect the context of our clinic, which is in Northern New England with limited racial and ethnic diversity.

**Conclusions:** The co-design process resulted in a simplified CKD Talk Topics intervention. These changes supported our goal: a patient-centered, structured agenda-setting tool to help people with advanced CKD and their care partners communicate priorities during nephrology visits. Feasibility testing is planned.

## Background

People with chronic diseases, including chronic kidney disease (CKD), who take an active role in self-management can have better health outcomes and improved quality of life.^1^ However, people with multiple health conditions, including those with CKD, can struggle with self-advocacy. In one study in the Veteran population, those with CKD asked fewer questions and received fewer answers than people with other serious illnesses.^2^ In this study, more than 90% of people with CKD wanted more information about their disease, but 40% said they did not ask their physician any questions, and nearly 50% said they could not ask informed questions due to their limited baseline knowledge of CKD and difficulty understanding clinician explanations.^2^ This sort of communication barrier can create dissatisfaction among patients and family members and knowledge gaps between patients and clinicians.^3–5^ A disproportionate number of people with CKD also live in rural areas where access to services and support for managing the complex physical and emotional demands of CKD are limited.^6^ Rural-residing people with CKD also experience fewer opportunities to advocate for themselves and meaningfully engage in their care.^3–5^

Patient-centered communication interventions for people with CKD could theoretically improve information exchange between patients and clinicians by supporting patients to fulfill a more active role in clinical encounters and by supporting clinicians in receptive or listening roles.^7^ And there is some evidence that skillful clinician communication can build trust between clinician and patient.^8–11^ A systematic review of randomized controlled trials from Lunardi and colleagues also identified a variety of patient-activation interventions for people with CKD, with documented positive effects on self-management and self-efficacy.^12^ The interventions were varied and included specially tailored support such as coaching, motivational interviewing, setting up individual care plans, goal-setting, problem-solving, culturally-tailored meetups, and confidence-building exercises.^12^ Similarly, a systematic review from Lopez-Vargas and colleagues identified more than two dozen educational interventions for people across all stages of CKD, although they were largely unidirectional information giving, and self-advocacy outcomes were not assessed.^13^ There is a call for more research and innovation concerning communication and coordination in CKD care.^14^

Given the challenges people with CKD have in asking their clinicians questions, there is a need for interventions that explicitly increase self-advocacy in the clinical visit. Collaborative agenda-setting is a patient-centered communication practice where clinicians work together with patients to “elicit, propose and organize” discussion topics for the visit, supporting patients in self-advocacy.^15–17^ Although there have been many agenda-setting interventional studies,^18–21^ we are not aware of any interventions specifically designed to support people with CKD generally or advanced CKD specifically.

Given the gap in agenda-setting interventions to increase self-advocacy among people with advanced CKD, we conducted a formative and participatory qualitative research study to develop a structured agenda-setting (SAS) tool (CKD Talk Topics).

## Materials and Methods

### Design

We conducted a participatory qualitative study to develop CKD Talk Topics using semi-structured interviews in the context of a larger parent study on advanced CKD. This work was conducted in a rural-serving Nephrology clinic in Northern New England. We used the Serious Illness Topics structured agenda-setting intervention as a model for CKD Talk Topics.^22^ SIT is a tool designed to support patient-centered visits for people with any serious illness.^22^ Serious illnesses are any diseases or conditions with a high expectation of mortality that either burden care partners or limit activities of daily function.^23^ Serious illnesses include but are not limited to advanced cancer, heart disease, kidney disease, Alzheimer’s, chronic obstructive pulmonary disease (COPD), and others.^24^ SIT has 8 plain-language discussion topics rooted in the most common concerns of people with serious illnesses; we developed it with participatory design approaches.^22^ SIT and derivative structured agenda-setting interventions are meant to be offered repeatedly at every clinic visit.^22,25^ An adaptation of SIT has been used in more than 3,000 visits as a part of routine care in the Dartmouth Hitchcock Medical Center’s (DHMC) outpatient palliative clinic.^25^

We report our results in line with the Consolidated criteria for Reporting Qualitative Research (COREQ).^26^ Our Community Advisory Board (CAB), which included three people living with kidney disease and two care partners, was engaged in this study from conception through dissemination.

### Participants

We purposively selected participants to obtain a broad range of roles (patient, clinician) and demographic characteristics. We also monitored balance across sex and socioeconomic status (measured by income and education). We included patients with advanced CKD not on dialysis, as their experiences are materially different from those on dialysis, and the patients on dialysis receive care in a separate facility with a separate workflow. We defined advanced CKD as stages 4 (estimated glomerular filtration rate (eGFR) 15-29 L/min/m2) and 5 (eGFR<15) at the time of screening.^27^ We required participants to be 18 or older, English-speaking, able to provide informed consent, and to be attending an outpatient visit in-person at the Dartmouth Hitchcock Medical Center’s (DHMC) Nephrology clinic. Additionally, we selected DHMC clinicians who regularly care for patients with advanced CKD in this clinic, including physicians, advanced practice providers, nurses, and support staff.

### Screening, recruitment and procedures

We identified eligible patients through electronic health record review ahead of appointments at the DHMC Nephrology clinic and communicated the time of appointments to the study team for recruitment. The study team confirmed eligibility with nurse clinicians (VJ, LG). For clinician participants, we received a list of eligible individuals directly from our Nephrology clinician collaborators, and we had pre-existing relationships with some of the participants (COREQ 6).

A researcher explained the study procedures, invited patients to participate in the study, and obtained informed consent from those who were eligible and willing to participate, and scheduled a one-on-one interview in person in a non-clinical area (COREQ 14). Interviews took anywhere from 45-90 minutes (COREQ 21). Field notes were completed by the interviewer immediately following the interview. We audio recorded interviews with participants’ permission and had the recordings professionally transcribed verbatim. One clinician asked us to take notes without recording the interview, due to regulatory matters related to their employment contract (COREQ 19-20). We compensated patient participants, but not accompanying care partners, with $25 physical gift cards and offered thank-you letters.

### Data collection instruments

We developed and piloted a semi-structured interview guide, with guidance from our CAB (COREQ 17). The guide had focused questions on current agenda-setting practices, expectations for structure and topics of an agenda-setting intervention, and implementation of that agenda-setting intervention in the Nephrology practice’s dedicated CKD clinic.^28,29^ This portion of the interview guide was informed by cognitive interview ‘think aloud’ techniques.^22,30,31^ The guide also explored the needs and experiences of people with CKD, with a particular focus on aspects of care that might be germane to patients living in rural areas such as extended distances traveling to appointments and difficulty procuring medications and supplies across large geographic distances.^32^ We analyzed this portion of the data in another manuscript, but have drawn on some of these findings in intervention development.^32^

We shared the SIT intervention as a model along with subsequent iterations of CKD Talk Topics during interviews and repeatedly with our CAB and solicited feedback on how the construct could be adapted for the advanced CKD-specific context.^33^ Additionally, the interview guide included an initial list of discussion topics based on a literature review about the experiences of people with advanced CKD.^34–38^ A version of the interview guide for patients is available in Appendix 1, and the clinician guide in Appendix 2.

In addition to the interviews, we administered a survey to each participant. The survey included standard demographic questions and Weiner’s validated Feasibility of Intervention Measure (FIM). To identify opportunities for increased feasibility, we monitored these scores, aiming for ≥3 on the 5-point scale.^39–41^ The survey for patients is available in Appendix 3, and the clinician survey in Appendix 4. We reported the characteristics of the interviewers and relationships with the participants (COREQ 1-8).

### Analysis

#### Framework and practical thematic analysis

For understanding participants’ feedback on both the CKD agenda-setting tool and implementation guide, we used Framework Analysis.^42,43^ Three researchers (AD, AZ, IK) read over the transcripts, identified relevant quotes, and developed summaries for each cell of a matrix, with field notes and memos as-needed (COREQ-20). The matrix was organized with key questions from the interview guide regarding tool adaptation as its columns (Appendix 5) (COREQ-25).^42,43^ Two independent researchers used the initial summaries to develop justified meta-summaries by column (CHS, EC). Facilitators (CHS, AS) led a discussion with the researchers (AD, AZ, IK) to confirm and refine the meta summaries. Those meta-summaries, both from patient, patient (plus care partner), and clinician interviews, informed the development of CKD Talk Topics. We cross-referenced these final meta-summaries against the raw data to check for accuracy. We iteratively revised our agenda-setting intervention between data collection and analysis rounds, conducting additional interviews as needed to be responsive to feedback (COREQ-18) (Figure 2). For CKD Talk Topics development, we considered data saturation reached once we were not receiving substantive suggestions for modification. We discussed data saturation in-between rounds of tool iteration (COREQ-11). Our Practical Thematic Analysis approach, which is a participatory approach to qualitative data analysis, is described elsewhere, and we have drawn on these findings during our intervention development.^32^ When feedback from interviewees conflicted, we used a feedback hierarchy protocol to make final design decisions. See **Figure 1**.

**Figure 1.**
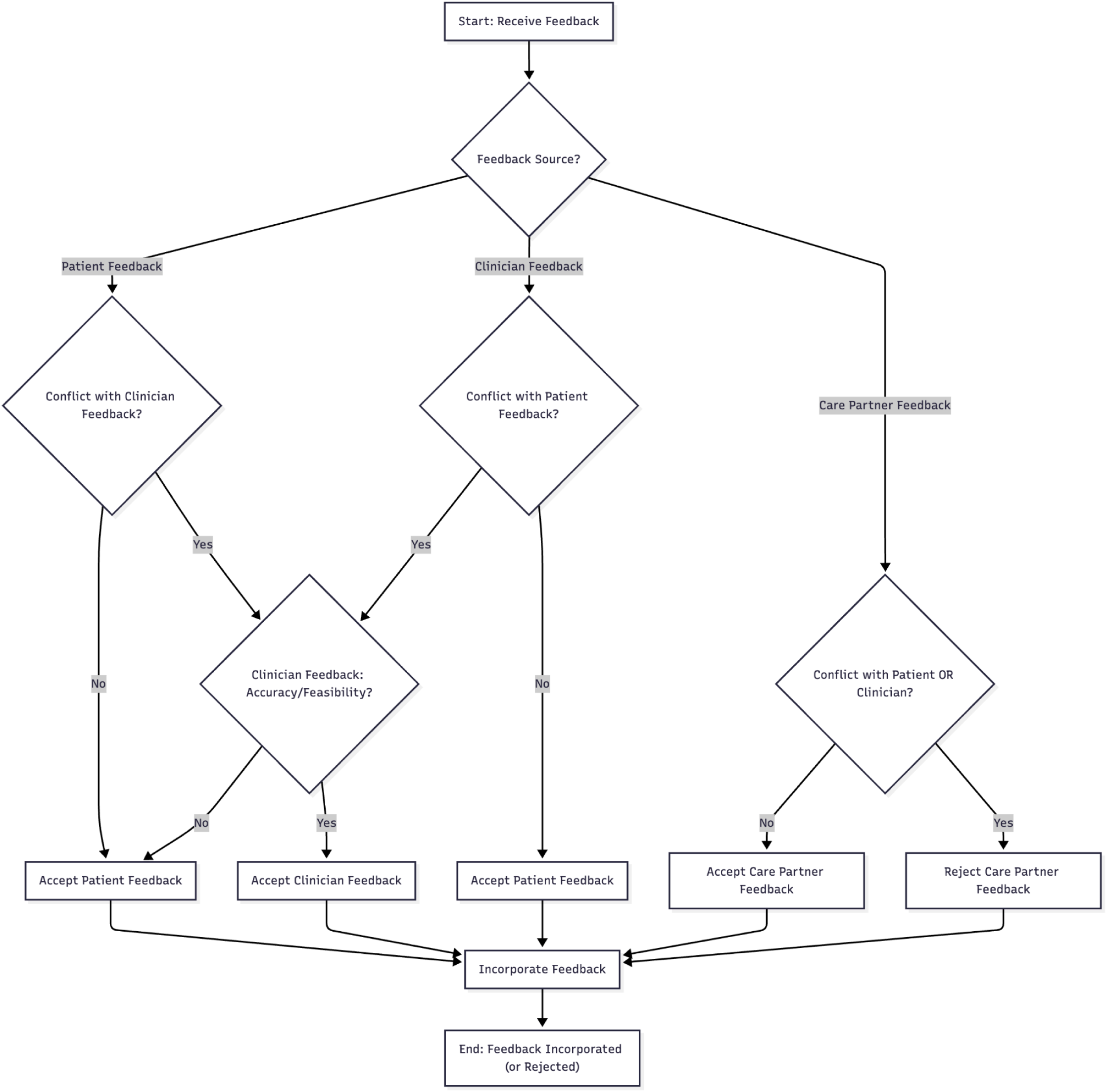
Patient, care partner, clinician feedback hierarchy. **Note:** We used Google GenAI Gemini 2.0 Flash and Mermaid AI to refine this model in Mermaid Chart.

**Figure 2.**
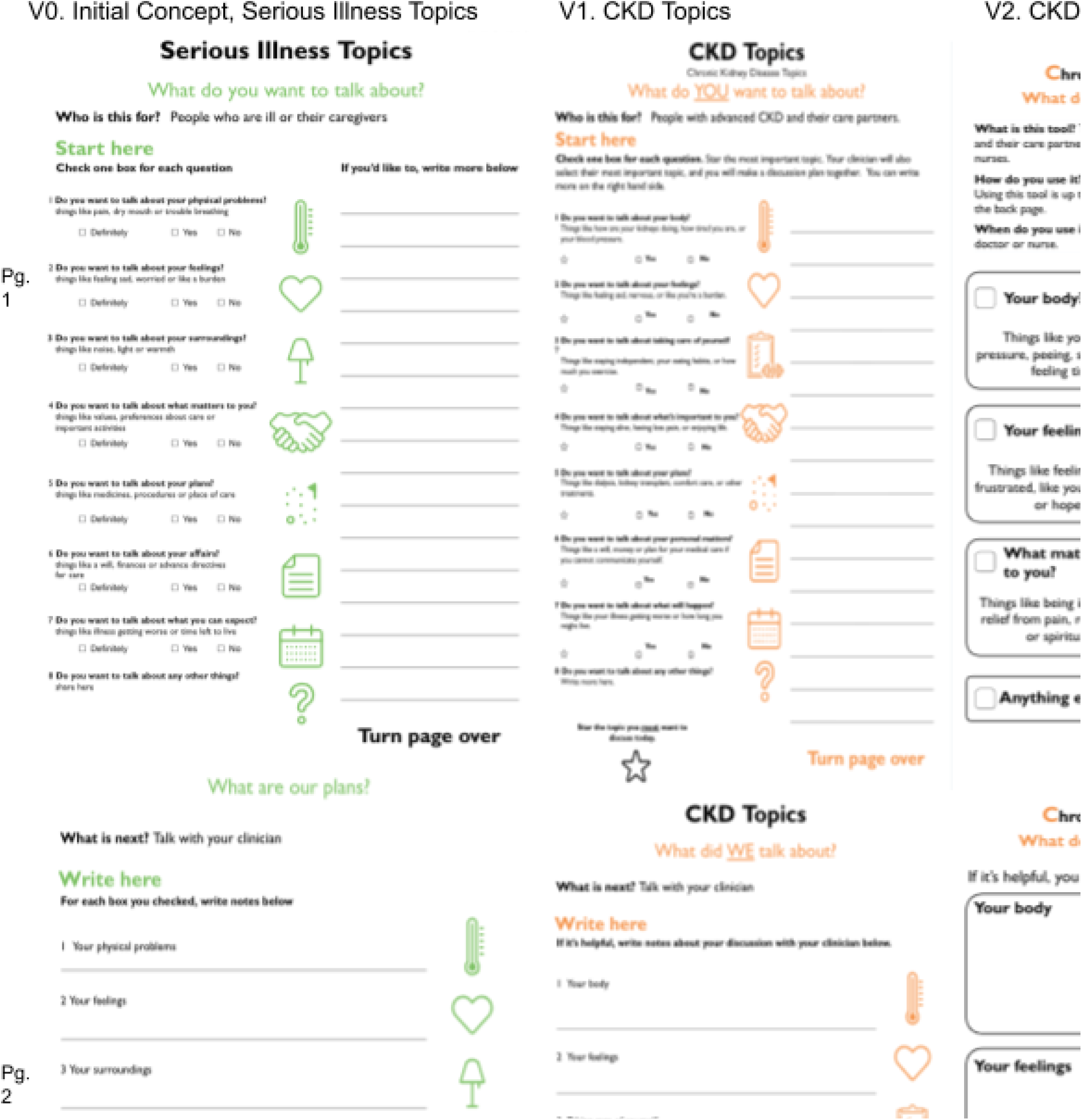
Iterative evolution of CKD Talk Topics structured agenda-setting intervention.

### Assessment of final intervention

We conducted Flesch-Kincaid and Linsear Write Grade Level assessment of CKD Talk Topics, and revised the language to achieve sufficiently accessible material. Flesch-Kincaid is often used to assess government forms and technical manuals, targeting a 9th grade reading level or below. Linsear is based on the average sentence length and how often complex words occur.^44^

## Results

### Characteristics of sample

#### Participant demographics

We interviewed 12 patients, four of their care partners, and eight clinicians. Among the patient participants, eight (67%) were male and four (33%) were female (**Table 1A**). Most were 55 years or older (n=10, 83%), reported no difficulty accessing healthcare (n=10, 83%), and had attained a high school diploma or equivalent or less (n=9, 75%). All participants identified as White or Caucasian (n=12, 100%). Our clinician interviews included three male (38%) and five female (63%) participants. Half of the clinician participants were physicians (n=4, 50%), had been in practice for 21 years or longer (n=4, 50%), and were 35-54 years old (n=4, 50%) (**Table 1B**). Most participants identified as White or Caucasian.

**Table 1A.** Characteristics of included participants (patients)*

|  | Number of patient participants<br>N=12 |
| --- | --- |
| **Age** |  |
| 35-54 years old | 2 |
| 55-74 years old | 6 |
| 75+ | 4 |
| **Gender** |  |
| Female | 4 |
| Male | 8 |
| **RUCA code** |  |
| 4-6 (Large rural) | 5 |
| 7-9 (Small rural) | 2 |
| 10 (Isolated rural) | 5 |
| **Healthcare access difficulty<sup>†</sup>** |  |
| No | 10 |
| Yes | 0 |
| **Education** |  |
| Less than high school | 3 |
| High school diploma or equivalent | 6 |
| Some college, Associate's or Bachelor's degree | 3 |
| **Insurance<sup>†</sup>** |  |
| Medicaid | 3 |
| Medicare | 4 |
| Private insurance | 4 |
| **Comfort with medical forms (health literacy)<sup>†</sup>** |  |
| No comfort/ little comfort | 4 |
| Quite a bit/ extremely comfortable | 7 |
| **Employment** |  |
| Full-time | 3 |
| Part-time | 2 |
| Unemployed | 2 |
| Retired | 5 |

\*Reproduced from Sanders et. al.<sup>32</sup>
† Missing data: healthcare access difficulty (n=2), insurance (n=1), health literacy (n=1)
<sup>1</sup>All participants in this sample identified as "Caucasian or White".
<sup>2</sup> We used Fisher's exact test given the small sample size
<sup>3</sup> Health literacy status was referring to the question: "How confident are you in filling out medical forms by yourself? From Chew, Morris and colleagues"<sup>68-70</sup>

**Table 1B.** Characteristics of included participants (clinicians)

|  | Total (N=8, %) |
| --- | --- |
| **Age** |  |
| 18-34 years old | 1 (13) |
| 35-54 years old | 5 (63) |
| 55-74 years old | 2 (25) |
| 75+ | 0 (0) |
| **Years in practice** |  |
| 1-10 years | 3 (38) |
| 11-20 years | 1 (13) |
| 21+ years | 4 (50) |
| **Clinical role** |  |
| Physician (MD/DO) | 4 (50) |
| Nurse practitioner | 1 (13) |
| Registered nurse | 2 (25) |
| Other | 1 (13) |
| **Race** |  |
| White or Caucasian | 6 (75) |
| Asian | 1 (13) |
| Other | 1 (13) |

**Table 3.** Themes and associated intervention changes.

**Table 3a.**
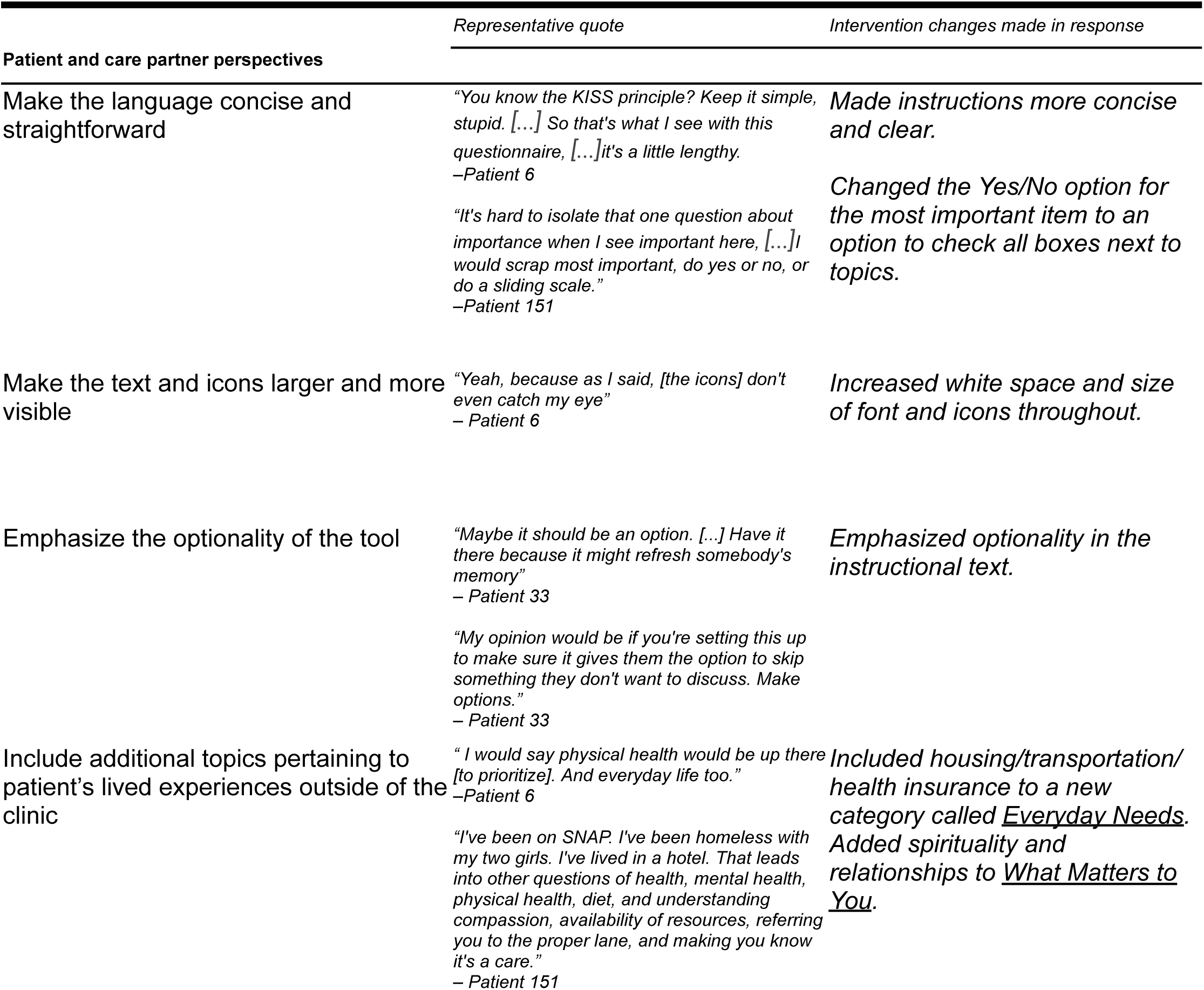
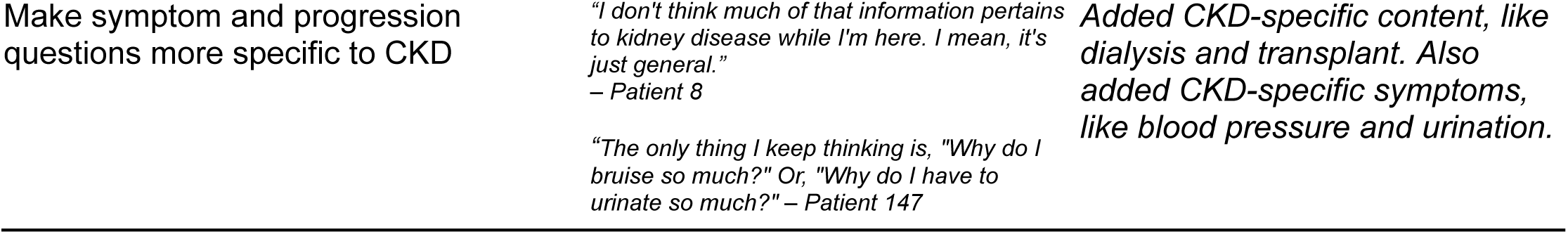
Patient and care partner perspectives.

**Table 3b.**
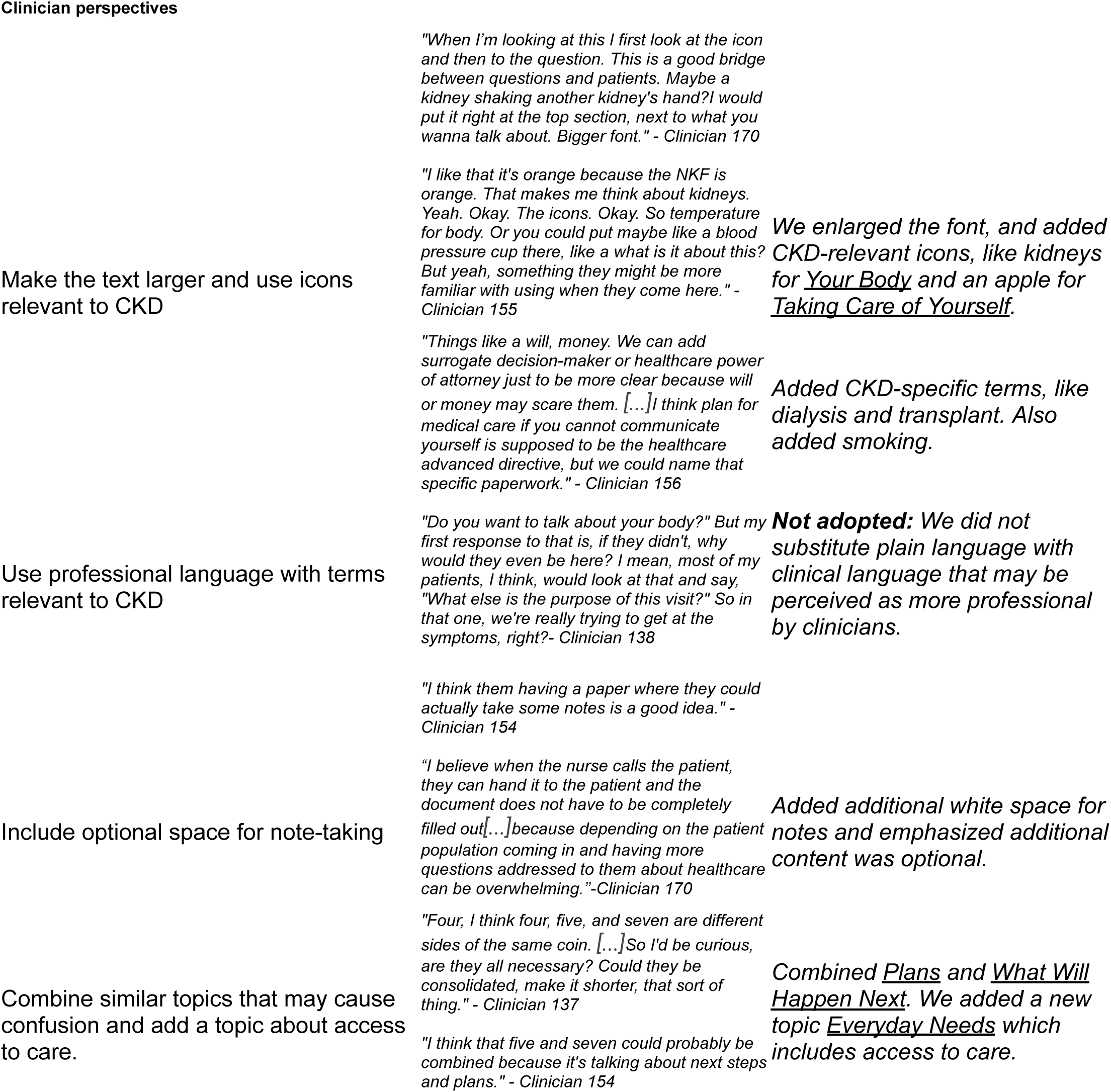
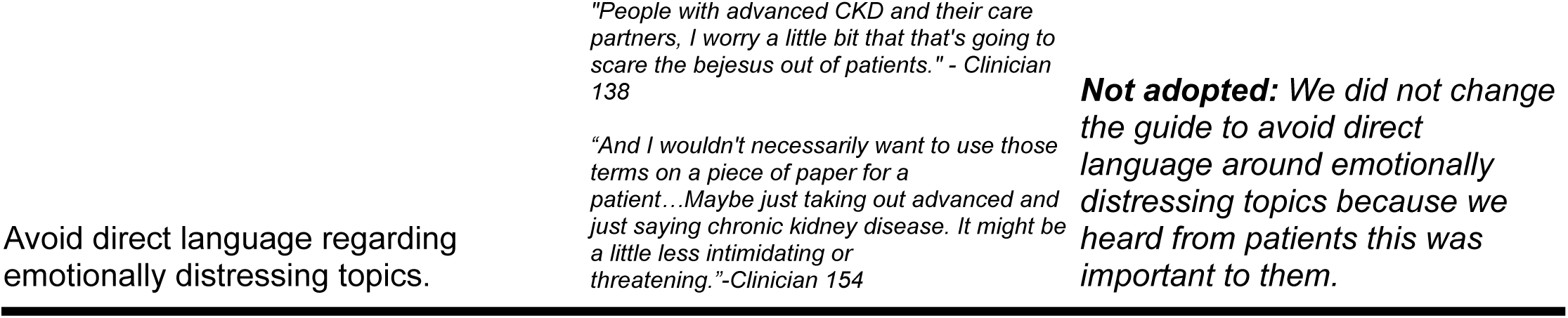
Clinician perspectives.

### Lived experiences changes

In addition to the discrete themes described above, through our PTA process, we also identified themes about the lived experiences and preferences of rural-residing patients and care partners. These findings are described in detail elsewhere, but the themes were as follows:

1. Physical symptoms and lifestyle changes were common but often attributed to comorbid conditions rather than CKD.
2. Logistical challenges of rural CKD care were pervasive and often normalized.
3. Disease progression and future treatments were sources of uncertainty and concern.
4. Clinical conversations often center on lab results and medications, leaving emotional concerns unaddressed.

Given the rurality theme, we emphasized transportation issues in CKD Talk Topics. We also signaled that feeling apprehensive about the course of CKD was normal with the topics “Feelings” and “What Happens Next” in response to the Future theme. We retained and elaborated on the “Feelings” topic in response to the Communication theme and added CKD-specific symptoms in response to the Symptom theme.

### Primary analysis findings

#### Patient and care partner interview findings

Overall, patients and care partners expressed appreciation for the CKD Talk Topics SAS and the general concept of SAS. One participant wondered why this was not already the standard of care in practice, stating, “something like this could be an overall thing across other specialties” (patient participant 151). We heard from participants that visits are often dominated by discussions about lab values, and it can be challenging to discuss bigger-picture items, like disease progression and emotions in the context of the clinical visit; “you need some sort of psychological safety to talk about some of these things” (patient participant 151). Participants believed an agenda-setting intervention could open the door to these types of discussions.

We identified five main themes in patients’ and care partners’ feedback on the design of our agenda-setting intervention:

1. Make the language concise and straightforward
2. Make the text and icons larger and more visible
3. Include additional topics pertaining to patient’s lived experiences outside of the clinic
4. Make symptom and disease progression questions more specific to CKD
5. Make sure the tool and the elements of the tool are clearly optional

#### Make the language concise and straightforward

Most patients and their care partners appreciated the concise and straightforward language in the initial SIT intervention and subsequent CKD Talk Topics iterations, especially around topics related to disease progression. However, some felt that the language used in the intervention was overly wordy and unclear, emphasizing that shorter, more direct wording would make the tool easier to navigate and less burdensome to complete. In an initial draft of the intervention, participants were prompted to select which topic they felt was “most important” to discuss, which some found difficult, as they perceived most topics as important to discuss and had difficulty prioritizing.

In response to this feedback, we worked to make the instructions more concise and clear. We also changed the option boxes that were previously asking “Yes/No” in response to “Most important?” to a simple open checkbox next to each topic. The instructions were adapted to say “Check the topic boxes that you want to talk about” instead of “Check one box for each question”. This reduces the impression that each topic is a question in need of a response, with the intent of decreasing respondent burden.

#### Make the text and icons larger and more visible

While most participants found the visual layout of the intervention easy-to-follow, some felt there was room for improvement. Specifically, some patients mentioned the need for larger font sizes and more visible icons to support readability, particularly for older adults or those completing the tool on tablets or phones.

In response to this feedback, we made room for more white space and made the font bigger. These changes made the icons easier to see.

#### Include additional topics pertaining to patients’ lived experiences outside of the clinic

Most patients and care partners responded positively to the intervention’s topics, noting that the issues they wanted to discuss included symptoms, disease progression, and daily management. However, many patients felt that the current tool did not capture the broader context of living with a serious illness like CKD, including topics that can be particularly salient with rural living, like housing, transportation, and family or other social support systems, including spirituality. For instance, we learned from the thematic analysis of lived experiences, many CKD patients are often dependent on support from others for transportation to clinical visits, and this can be especially challenging in rural areas. Patients and care partners felt that broadening the scope of the agenda-setting intervention to address these concerns would allow for more comprehensive conversations with their clinical teams about topics important for their disease management.

In response to this feedback, we changed “Personal Matters” to “Everyday Needs” and included housing, transportation, money, and health insurance while keeping goals of care/advanced planning constructs in “What Matters to You.” We also added spirituality and relationships under the “What Matters to You” topic.

#### Make symptom and disease progression questions more specific to CKD

Patients wanted to talk about specific experiences they had around CKD disease progression and symptoms. Instead of asking “Do you want to talk about your plans?” which patients found to be too vague, they preferred more straightforward, direct and specific language around particular treatment options and decisions, such as dialysis and transplant. One patient suggested having a prompt for symptoms around urinary changes, noting that this change would provide more clarity than the original question, “Do you want to talk about how your kidneys are doing?”

In response to this feedback, we added both urination (which we referred to as ‘peeing’ for plain language in the tool) and lab results to the “Your Body” construct. We also changed the construct of “Your Plans” to “What Happens Next” and included CKD-related advanced treatment prompts such as dialysis and transplant.

#### Make sure the tool and the elements of the tool are clearly optional

Participants expressed that while the intervention may be useful in guiding conversations with clinicians, it should remain optional rather than mandatory at every visit, so patients do not feel pressured or burdened to use it. There was concern that the tool may become less meaningful if repeated use among participants who visit their clinicians frequently was required, or even emotionally distressing, especially if it raises difficult topics. Participants felt that having the flexibility to use and complete elements of the intervention only when they felt the need to do so resolved this issue. The participants also shared that choices around using the tool would allow them to control how and when they used it, skipping questions or discussion topics that they did not wish to discuss, and preserving their autonomy and emotional comfort. Participants expressed that wording emphasizing how and whether patients used the tool was their choice would help them feel empowered to guide the discussion based on their own comfort and needs.

In response to this, we emphasized that the tool was optional in the instructions and in the checkbox option for indicating topics of choice. To underscore the optional nature of the intervention without deterring patients from completing it, we used language that emphasizes patient choice and flexibility. Phrases such as “suggested topics,” and “using the tool is up to you” were included to convey that engagement with the tool is voluntary. On the back page of the tool, we again emphasized using the open space to write notes was optional, not required.

### Clinician interview findings

We identified five main themes in clinicians’ feedback on our agenda-setting intervention:

1. Make the text larger and use icons relevant to CKD
2. Use professional language with terms relevant to CKD
3. Include optional space for note-taking
4. Combine similar topics that may cause confusion and add a topic about access to care
5. Avoid direct language regarding emotionally distressing topics

Findings from clinician interviews were incorporated into the tool redesign when not in direct conflict with patient and care partner preferences, as per our methodological prioritization of patient feedback (Figure 1).

#### Make the text larger and use icons relevant to CKD

Much like the patients and care partners, clinicians stressed the importance of readability and accessibility, noting that most members of the patient population were older and/or already experiencing a significant cognitive burden associated with managing their disease and navigating health systems. For visual appeal and relevance, clinicians suggested updating icons more familiar to patients with CKD, like using a blood pressure cuff instead of a thermometer to portray symptoms. We updated the intervention’s font size and icons in response to this feedback.

#### Use professional language with terms relevant to CKD

Clinicians suggested that examples under suggested topics use more professional terminology, and some recommended using terms more closely linked to CKD disease progression and treatment, such as urination, blood pressure, smoking or alcohol use, dialysis, and transplant. Some clinicians stated that “quality of life” should be used in place of “staying alive” and that pain control should be explicitly asked about.

In response to this feedback, we included urination (“peeing” in plain language, also in response to patients’ and care partners’ thoughts on language) under “Your Body.” Regarding the feedback on asking about “quality of life”, we reworked “What Matters to You” to include advanced care planning constructs without directly asking about “quality of life,” which as a term may be confusing to patients. Given our feedback hierarchy, whenever relevant, we elected to keep our language plain and understandable to patients, instead of using more technical terms. This is also consistent with evidence that patients prefer plain, understandable language.^45^

#### Include optional space for note-taking

Much like patients and care partners, clinicians emphasized the importance of not having any content or structure in the intervention that would suggest it was mandatory for patients, as they did not want them to feel pressured or overwhelmed. Clinicians also thought additional space for writing and context would be helpful for patients and also informative for clinicians. In response to this feedback, we emphasized the open space and optionality of the notes space on the back.

#### Combine similar topics that may cause confusion and add a topic about access to care

There was a general consensus among clinicians that overlapping topics should be combined to avoid confusion and clarify intent. In particular,the topics related to planning for the future, advanced planning and personal matters, and patient values about quality of life as CKD progresses were perceived as conceptually similar. Clinicians suggested adding topics like access to care, education about CKD, and barriers to care.

This influenced the addition of a new additional topic about everyday needs encompassing transportation, money, housing, and insurance. We also combined elements of four overlapping items pertaining to advanced planning and the future into two consolidated topics. We also added the construct of “Everyday Needs” to include socioeconomic concerns that may impact patients’ experience of CKD and access to treatment, like housing and transportation.

#### Avoid direct language regarding emotionally distressing topics

Some clinicians expressed concern with using direct language about potentially emotionally distressing topics. This was often connected to concerns regarding how to address challenging topics in a clinical visit, particularly those perceived by clinicians as outside of the Nephrology clinic’s scope, like mental/psychological wellbeing and/or palliative or end-of-life advanced planning.

Given evidence that patients want clinicians to be direct about their diseases getting worse and often find euphemisms and medical jargon confusing, we elected to leave this language as-is and not make the changes clinicians suggested about avoiding language around direct or emotionally distressing topics. This is also consistent with our feedback hierarchy^46^ and medical consensus guidelines.^47^

To be supportive of clinician concerns about feeling certain topics are outside their scope, we are developing a separate resource about how to respond to emotion, which will be described elsewhere. It will include resources and referrals for topics that are out of a clinician’s comfort area as they use CKD Talk Topics.

### Final CKD Talk Topics intervention

The final CKD Talk Topics SAS (Appendix 6) has six discrete topics based on what we know people with CKD care about discussing with clinicians. These topics include: “Your Body,” “Your Feelings,” “What Matters to You,” “Taking Care of Yourself,” “What Happens Next”, and “Everyday Needs.” It also has an additional topic area for anything else that falls outside the first six topics. The first page has instructions, along with checkable topics supplemented with descriptive icons. The second page has additional space to elaborate on concerns or notes on the discussion with the clinician. The final document has a Flesch-Kincaid 5th-grade reading level and a Linsear Write Grade Level of 3rd-grade reading level.^44^

## Discussion

### Summary of results

We developed a brief, simple, SAS intervention called CKD Talk Topics. CKD Talk Topics has six topical discussion areas, plain language, and descriptive icons. We designed the tool to support collaborative agenda-setting between patients, their care partners, and their clinicians. The content and structure of the tool were informed by patient and care partner preferences for clarity, simplicity, and CKD-specific material.

Clinicians also wanted CKD-specific material, but preferred more technical and medical language.

### Results in context

To our knowledge, this is the first agenda-setting intervention specifically made for people with kidney disease. We know of some agenda-setting interventions for other serious illness contexts. Notably, the SIT agenda-setting intervention, from which CKD Talk Topics was adapted, is for people with serious illnesses, largely those in the final months or years of life.^22^ There are a variety of agenda-setting interventions for other serious illnesses and chronic disease contexts, like major depressive disorders,^48^ respiratory diseases,^19^ diabetes,^18,49^ hypertension,^50^ and oncology.^20,21,51^ There are also many generalized agenda-setting interventions for primary care contexts.^50,52^ Although there are no CKD-specific agenda-setting interventions, there are other patient-centered interventions specifically for people with CKD, including advanced CKD, like decision aids^53–55^ and advanced care planning interventions.^56^ Reviews, mentioned earlier, from Lunardi and Lopez-Vargas show promising benefits from a broad range of patient activation and communication interventions, and CKD-specific interventions, respectively, but it remains to be seen whether CKD Talk Topics improves patient-centered communication outcomes.^12,13^ We plan to test this in a stepped-wedge randomized clinical trial.

Some of the phenomena we uncovered were consistent with other literature about people with advanced CKD. For instance, in a fieldwork based study of clinical visits, Kang Stenfors-Hayes found that patients and clinicians had different mental models about goals for care, and there were notable “tensions and gaps in understanding.”^57^ Our finding that patients wanted space to talk about disease progression and emotional concerns, while clinicians were uncomfortable with providing that space is reminiscent of their findings.

Some of the discrepancies between patient and clinician perspectives about what is important to discuss in the clinical encounter may support what we already know about the multifaceted and complex power dynamics between patients and clinicians in the clinical encounter. ^58^ For instance, although the emotional impacts of CKD are well-known, clinicians in our study did not want or did not feel comfortable creating space for such discussions, preferring the clinician-directed implicit agenda-setting that occurs currently.^59^ These tensions between patient and care partner expectations and preferences, and those of physicians are not unique to agenda-setting contexts; there are growing perceptions that patient expectations overall have changed as healthcare has become more consumeristic.^60,61^ Simultaneously, patients’ rights movements have exposed fault lines between patient expectations in healthcare writ large, with arguments for more patient protections in insurance, in dealings with healthcare systems and more.^62^

### Implications

Our findings through the development of this intervention demonstrate that patients are interested in discussing the progression of their disease with their clinicians, but may not know how to start those conversations. Clinicians, however, are less comfortable discussing emotion with their advanced CKD patients. Without purposeful disruption, this power dynamic is unlikely to change.^58,63^ Some clinicians are likely to be more open to reflection and suggestions about dynamics of power and power sharing in the clinical encounter.^58^ To mitigate this discrepancy, nephrology leaders may consider offering training in serious illness conversations, like the Serious Illness Conversation Guide, or NephroTalk, which is a Nephrology-specific serious illness training program.^64,65^ These types of trainings may build serious illness conversation and emotional conversation literacy among clinicians. Relatedly, it is well-known that most physicians do not receive significant training in how to respond to patients’ emotions or their own emotions during clinical encounters, and this was another notable discrepancy between patient and clinician perspectives, so these types of trainings may enhance physicians’ ability to respond to these scenarios.^66^ An agenda-setting tool is only an entry point for these personalized discussions, but clinicians will need more support to feel comfortable talking with their patients about complex feelings about their disease. These types of training may be beneficial beyond nephrology as well.

Our findings also have implications for other researchers developing interventions; clinician and patient feedback can be in direct conflict, and having a clear way of managing these conflicting preferences is critical. Our feedback hierarchy has been a helpful guide for our team in managing these conflicts when they arise.

### Limitations

This study was unique to the rural-serving clinic in which it was performed, and the intervention may need to be confirmed as appropriate for other contexts. The population was white, and tended to be older than the CKD population at large. Although we had a small qualitative sample, this is within standard guidelines for qualitative research, and data saturation was reached, suggesting our sample was sufficient.^67^ One notable limitation of this study is that we were only able to speak with people who came to the clinic on-site, as opposed to those who had telehealth visits. It is possible that the needs and experiences of those who do video or phone visits are different from those for on-site patients. An upcoming open feasibility pilot is planned, and we will use this opportunity to assess appropriateness for telehealth.

## Conclusion

We developed a simple, patient-centered SAS that will require further feasibility and efficacy testing.

## Supporting information

Supplemental Quotes

Supplemental COREQ

## Data Availability

The data from this study are not publicly available because the interview data cannot be fully de-identified.

## Acknowledgements

We acknowledge Scott G O’Rourke for his support recruiting patients for this study. We would also like to sincerely thank all the patients, care partners and clinicians who participated and lended their perspectives.

## References

1. Ma B, Wang H, Jia Y, et al. Systematic review and network meta-analysis of the comparative effectiveness of self-management support strategies for patients with chronic kidney disease. Worldviews Evid Based Nurs. 2025;22(6):e70090. doi:10.1111/wvn.70090

2. Lederer S, Fischer MJ, Gordon HS, Wadhwa A, Popli S, Gordon EJ. Barriers to effective communication between veterans with chronic kidney disease and their healthcare providers: Table 1. Clin Kidney J. 2015;8(6):766–771. doi:10.1093/ckj/sfv079

3. Flythe JE, Karlsson N, Sundgren A, et al. Development of a preliminary conceptual model of the patient experience of chronic kidney disease: a targeted literature review and analysis. BMC Nephrol. 2021;22(1):233. doi:10.1186/s12882-021-02440-9

4. Almutary H, Bonner A, Douglas C. Symptom burden in chronic kidney disease: a review of recent literature. J Ren Care. 2013;39(3):140–150. doi:10.1111/j.1755-6686.2013.12022.x

5. Roberti J, Cummings A, Myall M, et al. Work of being an adult patient with chronic kidney disease: a systematic review of qualitative studies. BMJ Open. 2018;8(9):e023507. doi:10.1136/bmjopen-2018-023507

6. Wu YL, Wu YC, Akhmetzhanov AR, Wu MY, Lin YF, Lin CC. Urban-rural health disparity among patients with chronic kidney disease: a cross-sectional community-based study from 2012 to 2019. BMJ Open. 2024;14(7):e082959. doi:10.1136/bmjopen-2023-082959

7. O’Hare AM. Patient-centered care in renal medicine: Five strategies to meet the challenge. Am J Kidney Dis. 2018;71(5):732–736. doi:10.1053/j.ajkd.2017.11.022

8. O’Grady C, Dahm MR, Roger P, Yates L. Trust, talk and the dictaphone: Tracing the discursive accomplishment of trust in a surgical consultation. Discourse Soc. 2014;25(1):65–83. doi:10.1177/0957926513496354

9. Strokes N, Lloyd C, Girardin AL, et al. Can shared decision-making interventions increase trust/trustworthiness in the physician-patient encounter? A scoping review. Patient Educ Couns. 2025;135(108705):108705. doi:10.1016/j.pec.2025.108705

10. Jiang S, Wu Z, Zhang X, et al. How does patient-centered communication influence patient trust?: The roles of patient participation and patient preference. Patient Educ Couns. 2024;122(108161):108161. doi:10.1016/j.pec.2024.108161

11. Martin KD, Roter DL, Beach MC, Carson KA, Cooper LA. Physician communication behaviors and trust among black and white patients with hypertension. Med Care. 2013;51(2):151–157. doi:10.1097/MLR.0b013e31827632a2

12. Lunardi LE, Hill K, Xu Q, Le Leu R, Bennett PN. The effectiveness of patient activation interventions in adults with chronic kidney disease: A systematic review and meta-analysis. Worldviews Evid Based Nurs. 2023;20(3):238–258. doi:10.1111/wvn.12634

13. Lopez-Vargas PA, Tong A, Howell M, Craig JC. Educational interventions for patients with CKD: A systematic review. Am J Kidney Dis. 2016;68(3):353–370. doi:10.1053/j.ajkd.2016.01.022

14. Jurkovitz C, Dolman SF, MTAv, Archinal H. PARTNERS IN RESEARCH: Developing a patient-centered research agenda for chronic kidney disease. Dela J Public Health. 2017;3(1):24–29. doi:10.32481/djph.2017.03.004

15. Hood-Medland EA, White AEC, Kravitz RL, Henry SG. Agenda setting and visit openings in primary care visits involving patients taking opioids for chronic pain. BMC Fam Pract. 2021;22(1):4. doi:10.1186/s12875-020-01317-4

16. Rodriguez HP, Anastario MP, Frankel RM, et al. Can teaching agenda-setting skills to physicians improve clinical interaction quality? A controlled intervention. BMC Med Educ. 2008;8(1):3. doi:10.1186/1472-6920-8-3

17. Gobat N, Kinnersley P, Gregory JW, Robling M. What is agenda setting in the clinical encounter? Consensus from literature review and expert consultation. Patient Educ Couns. 2015;98(7):822–829. doi:10.1016/j.pec.2015.03.024

18. Robling M, McNamara R, Bennert K, et al. The effect of the Talking Diabetes consulting skills intervention on glycaemic control and quality of life in children with type 1 diabetes: cluster randomised controlled trial (DEPICTED study). BMJ. 2012;344:e2359. doi:10.1136/bmj.e2359

19. Early F, Everden AJ, O’Brien CM, Fagan PL, Fuld JP. Patient agenda setting in respiratory outpatients: A randomized controlled trial. Chron Respir Dis. 2015;12(4):347–356. doi:10.1177/1479972315598696

20. Wolff JL, Aufill J, Echavarria D, et al. Sharing in care: engaging care partners in the care and communication of breast cancer patients. Breast Cancer Res Treat. 2019;177(1):127–136. doi:10.1007/s10549-019-05306-9

21. Wolff JL, Roter DL, Boyd CM, et al. Patient-family agenda setting for primary care patients with cognitive impairment: The SAME page trial. J Gen Intern Med. 2018;33(9):1478–1486. doi:10.1007/s11606-018-4563-y

22. Saunders CH, Durand MA, Scalia P, et al. “It helps us say what’s important…” Developing Serious Illness Topics: A clinical visit agenda-setting tool. Patient Educ Couns. 2023;113:107764. doi:10.1016/j.pec.2023.107764

23. Kelley AS, Bollens-Lund E. Identifying the Population with Serious Illness: The “Denominator” Challenge. J Palliat Med. 2018;21(S2):S7–S16. doi:10.1089/jpm.2017.0548

24. Clinical Practice Guidelines. National Coalition for Hospice and Palliative Care. November 16, 2023. Accessed January 21, 2026. https://www.nationalcoalitionhpc.org/clinical-practice-guidelines/#:~:text=Improving%20Quality%20of%20Serious%20Illness,they%20live%20or%20receive%20care.

25. Wilson MM, Broglio K, Vergo MT, et al. An electronic pre-visit agenda-setting questionnaire in ambulatory palliative care is feasible and acceptable to patients, care partners, and clinicians: A mixed methods evaluation. Palliat Med. 2025;39(4):507–516. doi:10.1177/02692163251321327

26. Tong A, Sainsbury P, Craig J. Consolidated criteria for reporting qualitative research (COREQ): a 32-item checklist for interviews and focus groups. Int J Qual Health Care. 2007;19(6):349–357. doi:10.1093/intqhc/mzm042

27. Alcázar Arroyo R, Orte Martínez L, Otero González A. Advanced chronic kidney disease. Nefrologia. 2008;28 Suppl 3:3–6. https://pubmed.ncbi.nlm.nih.gov/19018731/

28. Damschroder LJ, Aron DC, Keith RE, Kirsh SR, Alexander JA, Lowery JC. Fostering implementation of health services research findings into practice: a consolidated framework for advancing implementation science. Implement Sci. 2009;4:50. doi:10.1186/1748-5908-4-50

29. Damschroder LJ, Reardon CM, Widerquist MAO, Lowery J. The updated Consolidated Framework for Implementation Research based on user feedback. Implement Sci. 2022;17(1):75. doi:10.1186/s13012-022-01245-0

30. Saunders CH, Durand MA, Scalia P, et al. User-Centered Design of the consideRATE Questions, a Measure of People’s Experiences When They Are Seriously Ill. J Pain Symptom Manage. 2021;61(3):555–565.e5. doi:10.1016/j.jpainsymman.2020.08.002

31. Saunders CH, Goldwag JL, Read JT, Durand MA, Elwyn G, Ivatury SJ. “Because Everybody is so Different”: a qualitative analysis of the lived experiences and information needs of rectal cancer survivors. BMJ Open. 2021;11(5):e043245. doi:10.1136/bmjopen-2020-043245

32. Sanders G, Kumar I, Dade A, et al. Normalized barriers and unaddressed concerns: A qualitative study of the lived experiences of adults living in rural areas with advanced chronic kidney disease. medRxiv. Published online July 10, 2026:2026.07.05.26356878. doi:10.64898/2026.07.05.26356878

33. Saunders CH, Durand MA, Scalia P, et al. “It helps us say what’s important…” Developing Serious Illness Topics: A clinical visit agenda-setting tool. Patient Educ Couns. 2023;113(107764):107764. doi:10.1016/j.pec.2023.107764

34. Baddour NA, Siew ED, Robinson-Cohen C, et al. Serious illness treatment preferences for older adults with advanced CKD. J Am Soc Nephrol. 2019;30(11):2252–2261. doi:10.1681/ASN.2019040385

35. Rosansky SJ, Schell J, Shega J, et al. Treatment decisions for older adults with advanced chronic kidney disease. BMC Nephrol. 2017;18(1):200. doi:10.1186/s12882-017-0617-3

36. Hazzan AD, Halinski C, Agoritsas S, Fishbane S, DeVita MV. Epidemiology and challenges to the management of advanced CKD. Adv Chronic Kidney Dis. 2016;23(4):217–221. doi:10.1053/j.ackd.2016.04.005

37. Teasdale EJ, Leydon G, Fraser S, Roderick P, Taal MW, Tonkin-Crine S. Patients’ experiences after CKD diagnosis: A meta-ethnographic study and systematic review. Am J Kidney Dis. 2017;70(5):656–665. doi:10.1053/j.ajkd.2017.05.019

38. House TR, Wightman A, Rosenberg AR, Sayre G, Abdel-Kader K, Wong SPY. Challenges to shared decision making about treatment of advanced CKD: A qualitative study of patients and clinicians. Am J Kidney Dis. 2022;79(5):657–666.e1. doi:10.1053/j.ajkd.2021.08.021

39. Leyenaar JK, Arakelyan M, Acquilano SC, et al. I-CARE: Feasibility, Acceptability, and Appropriateness of a Digital Health Intervention for Youth Experiencing Mental Health Boarding. J Adolesc Health. 2023;72(6):923–932. doi:10.1016/j.jadohealth.2023.01.015

40. Ellis S, Geana M, Griebling T, et al. Development, acceptability, appropriateness and appeal of a cancer clinical trials implementation intervention for rural- and minority-serving urology practices. Trials. 2019;20(1):578. doi:10.1186/s13063-019-3658-z

41. Weiner BJ, Lewis CC, Stanick C, et al. Psychometric assessment of three newly developed implementation outcome measures. Implement Sci. 2017;12(1):108. doi:10.1186/s13012-017-0635-3

42. Gale NK, Heath G, Cameron E, Rashid S, Redwood S. Using the framework method for the analysis of qualitative data in multi-disciplinary health research. BMC Medical Research Methodology. 2013;13(1):1–8. doi:10.1186/1471-2288-13-117

43. Goldsmith L. Using framework analysis in applied qualitative research. Qual Rep. 2021;26(6):2061–2076. doi:10.46743/2160-3715/2021.5011

44. Scott B. Readability Scoring System PLUS. Accessed November 11, 2025. https://readabilityformulas.com/readability-scoring-system.php

45. Allen KA, Charpentier V, Hendrickson MA, et al. Jargon be gone - patient preference in doctor communication. J Patient Exp. 2023;10:23743735231158942. doi:10.1177/23743735231158942

46. Barlet MH, Barks MC, Ubel PA, et al. Characterizing the language used to discuss death in family meetings for critically ill infants. JAMA Netw Open. 2022;5(10):e2233722. doi:10.1001/jamanetworkopen.2022.33722

47. Olsson MM, Windsor C, Chambers S, Green TL. A scoping review of end-of-life communication in international palliative care guidelines for acute care settings. J Pain Symptom Manage. 2021;62(2):425–437.e2. doi:10.1016/j.jpainsymman.2020.11.032

48. Deuter M, Martinez C, Preikschat B, et al. Effectiveness and Clinical Usefulness of Electronic Agenda-setting in Psychiatric Practices: A South Texas Psychiatric PBRN Study. J Psychiatr Pract. 2023;29(1):31–37. doi:10.1097/PRA.0000000000000686

49. Munch L, Stensgaard S, Feinberg MB, Elwyn G, Lomborg K. Evaluating the effect of Conversation Cards on agenda-setting in annual diabetes status visits: A multi-method study. Patient Educ Couns. 2024;119(108084):108084. doi:10.1016/j.pec.2023.108084

50. O’Malley PG, Jackson JL, Becher D, Hanson J, Lee JK, Grace KA. Tool to improve patient-provider interactions in adult primary care: Randomized controlled pilot study. Can Fam Physician. 2022;68(2):e49–e58. doi:10.46747/cfp.6802e49

51. Sepucha KR, Belkora JK, Mutchnick S, Esserman LJ. Consultation planning to help breast cancer patients prepare for medical consultations: effect on communication and satisfaction for patients and physicians. J Clin Oncol. 2002;20(11):2695–2700. doi:10.1200/JCO.2002.10.068

52. Kowalski CP, McQuillan DB, Chawla N, et al. “The Hand on the doorknob”: Visit agenda setting by complex patients and their primary care physicians. J Am Board Fam Med. 2018;31(1):29–37. doi:10.3122/jabfm.2018.01.170167

53. Fortnum D, Smolonogov T, Walker R, Kairaitis L, Pugh D. “My kidneys, my choice, decision aid”: supporting shared decision making: “ my kidneys my choice” decision aid. J Ren Care. 2015;41(2):81–87. doi:10.1111/jorc.12100

54. Ho YF, Kuo HL, Yang KL, Su JM. Development and usability testing of an interactive Patient-Decision-Aid App for patients with advanced chronic kidney disease. Int J Med Inform. 2025;201(105947):105947. doi:10.1016/j.ijmedinf.2025.105947

55. Fortnum D, Grennan K, Smolonogov T. End-stage kidney disease patient evaluation of the Australian “My Kidneys, My Choice” decision aid. Clin Kidney J. 2015;8(4):469–475. doi:10.1093/ckj/sfv050

56. Chan CH, Conley M, Reeves MM, Campbell KL, Kelly JT. Evaluating the impact of goal setting on improving diet quality in chronic kidney disease. Front Nutr. 2021;8:627753. doi:10.3389/fnut.2021.627753

57. Kang H, Stenfors-Hayes T. Feeling well and having good numbers: Renal patients’ encounter with clinical uncertainties and the responsibility to “live well”: Renal patients’ encounter with clinical uncertainties and the responsibility to “live well.” Qual Health Res. 2016;26(12):1591–1602. doi:10.1177/1049732315591484

58. Nimmon L, Stenfors-Hayes T. The “Handling” of power in the physician-patient encounter: perceptions from experienced physicians. BMC Med Educ. 2016;16(1):114. doi:10.1186/s12909-016-0634-0

59. O’Hare AM, Richards C, Szarka J, et al. Emotional impact of illness and care on patients with advanced kidney disease. Clin J Am Soc Nephrol. 2018;13(7):1022–1029. doi:10.2215/CJN.14261217

60. Shrank WH. Primary care practice transformation and the rise of consumerism. J Gen Intern Med. 2017;32(4):387–391. doi:10.1007/s11606-016-3946-1

61. The empowered patient: Consumerism in American medicine. AMA J Ethics. 2013;15(11):923–925. doi:10.1001/virtualmentor.2013.15.11.fred1-1311

62. Altman D. The Semi-Sad State of Consumer Protection In Health Care. KFF. January 7, 2025. Accessed February 5, 2026. https://www.kff.org/from-drew-altman/the-semi-sad-state-of-consumer-protection-in-health-care/

63. Emmerich N. Some relevant concepts. In: SpringerBriefs in Ethics. Springer International Publishing; 2013:1–20. doi:10.1007/978-3-319-00485-3_1

64. Bernacki R, Paladino J, Neville BA, et al. Effect of the Serious Illness Care Program in Outpatient Oncology: A Cluster Randomized Clinical Trial. JAMA Intern Med. 2019;179(6):751–759. doi:10.1001/jamainternmed.2019.0077

65. Cohen RA, Bursic A, Chan E, Norman MK, Arnold RM, Schell JO. NephroTalk multimodal conservative care curriculum for nephrology fellows. Clin J Am Soc Nephrol. 2021;16(6):972–979. doi:10.2215/CJN.11770720

66. Schwartz R, Osterberg LG, Hall JA. Physicians, emotion, and the clinical encounter: A survey of physicians’ experiences. Patient Educ Couns. 2022;105(7):2299–2306. doi:10.1016/j.pec.2022.03.001

67. Guest G, Bunce A, Johnson L. How Many Interviews Are Enough? Field methods. 2006;18(1):59–82. doi:10.1177/1525822X05279903

68. Chew LD, Bradley KA, Boyko EJ. Brief questions to identify patients with inadequate health literacy. Fam Med. 2004;36(8):588–594. https://www.ncbi.nlm.nih.gov/pubmed/15343421

69. Chew LD, Griffin JM, Partin MR, et al. Validation of screening questions for limited health literacy in a large VA outpatient population. J Gen Intern Med. 2008;23(5):561–566. doi:10.1007/s11606-008-0520-5

70. Morris NS, MacLean CD, Chew LD, Littenberg B. The Single Item Literacy Screener: evaluation of a brief instrument to identify limited reading ability. BMC Fam Pract. 2006;7:21. doi:10.1186/1471-2296-7-21

