## Supplemental Quotes for "Co-designing CKD Talk Topics, a structured agenda-setting intervention for people with chronic kidney disease: “Keep It Simple, Stupid”"

### Patient Interviews

**Quote list**

1. ***To make the language concise and straightforward***

Like I was telling you about **being straightforward is the way I like it to the point.** If I'm going to die next week, tell me. - Patient 30

I just want to you know I'm a basic simple guy. I don't need a lot of whipped cream. There's an issue. If I have a sickness, I want to be told I have a sickness, and I want to be told the facts. I want to be upfront. You know I don't want to sit back and listen to somebody talk to me for an hour and really not tell me anything. - Patient 8

But I like things you know the KISS principle. Keep it simple, stupid. All right? So that's what I see with this questionnaire, is that it's a little lengthy. Knowing and having worked with thousands of students over time, wow there's going to be certain things that they want to fill out, and some are not going to want to fill it out at all. - Patient 6

2. ***To make the text and icons larger and more visible***

“I just don't understand the thermometer [symbol], the heart [symbol]. I mean, you're asking questions. A lampshade matters to you. A handshake. The symbolism. I don't know if that's necessary.” Patient 6

“And then you know the graphics eliminate the graphics. They just do nothing for me. Yeah. And do you agree with them on the graphics? Yeah, because as I said, they don't even catch my eye.” Patient 6

“I'm not crazy about the icons, but But you understand what the But I understand. So are you suggesting to maybe use like what do you mean? What do you mean like you're not crazy about the icons? Oh, this is a thermometer. You're asking that it's on physical problems. Okay. So maybe you're suggesting different icons. Yeah, just different icons. I don't know. They don't make sense to me. Feelings, I get the heart. What could be changed or improved? I think better icons. Better icons.” Patient 33

3. ***To include additional topics pertaining to patient’s lived experiences outside of the clinic***

That covers a wide variety of topics. So you're liking the topics yeah for this group, this different group of patients. That'll cover your whole life. Yeah” Patient 30

4. ***To make symptom and progression questions more specific to CKD***

**“**And I don't think much of that information pertains to kidney disease while I'm here. I mean, it's just general.“ Patient 8

“Gosh, I would say all of the above. And based on our conversation today, it seems like that's something that's very important that you talk about during your visit, specifically around fatigue.” Patient 143

“So it really depends on what stage you're in. And this would be something that you would want to discuss later on or more like early? I think it would be subject for later.” Patient 33

“And the next question we have is, do you want to talk about taking care of yourself? So things like staying independent, your eating habits, or how much you exercise? Oh, yes. I always want to talk about that.” Patient 143

5. ***To emphasize the optionality of the tool***

“Maybe it should be an option. Option. You know Have it there because it might refresh somebody's memory” – Pt 26

“And if it's not, just that they don't have to fill out or something. Do you know what I mean? And I think the patients would feel more like they're involved because they're not just given this form like, "Here, fill this out." They're getting input first to see if they want to fill it out.” – Pt 31

“My opinion would be if you're setting this up to **make sure it gives them the option to skip something they don't want to discuss. Make options.** So here what we have for options is not required.” – Pt 33

“How helpful would it be? I think it would be there again. Helpful in the beginning, I think. It really depends on the individual and what the circumstances are.” – Pt 110

“But for somebody just I'm having kidney problems, what is it, a urinary tract infection, or is it I don't want to read all this stuff.” – Pt 110
