## Supplemental COREQ for "Co-designing CKD Talk Topics, a structured agenda-setting intervention for people with chronic kidney disease: “Keep It Simple, Stupid”"

### COREQ (COnsolidated criteria for REporting Qualitative research) Checklist

A checklist of items that should be included in reports of qualitative research. You must report the page number in your manuscript where you consider each of the items listed in this checklist. If you have not included this information, either revise your manuscript accordingly before submitting or note N/A.

| Topic | Item No. | Descriptions | Reported on Page No. |
|---|---|---|---|
| **Domain 1: Research team** | | | |
| Interviewer /facilitator | 1 | AD, AS, GS, JN, and CHS | Page 5 |
| Credentials | 2 | MPP, MD candidate, PhD student/MSc, MPH, PhD | Page 5 |
| Occupation | 3 | Research associate, MD candidate, PhD student, clinical research coordinator, health services researcher | Page 5 |
| Gender | 4 | Predominantly female (as identified in Procedures) | Page 5 |
| Experience and training | 5 | Participatory qualitative methods, palliative care, social research | Page 5 |
| **Relationship with participants** | | | |
| Relationshi p established | 6 | Not reported | N/A |
| Participant knowledge | 7 | Informed of study goals at recruitment | Page 5 |
| Interviewer characteris tics | 8 | Predominantly White; shared cultural context noted | Page 5 |
| **Domain 2: Study design** | | | |
| Methodolo gical orientation | 9 | Practical Thematic Analysis (PTA); Framework Analysis | Page 6 |
| Sampling | 10 | Purposive sampling | Page 5 |
| Method of approach | 11 | On-site recruitment (clinic visits) | Page 5 |

| | | | |
|---|---|---|---|
| Sample size | 12 | 12 patients, 4 care partners | Page 6 |
| Non-participation | 13 | 6 patients lost to follow-up | Page 5 |
| Setting of data collection | 14 | Northern New England Nephrology clinic | Page 5 |
| Presence of non-participants | 15 | Researchers and participants (some joint with care partners) | Page 5 |
| Description of sample | 16 | Demographics and metrics | Page 6 |
| Interview guide | 17 | Developed and piloted with CAB | Page 5 |
| Repeat interviews | 18 | No repeat interviews | Page 5 |
| Audio/visual recording | 19 | Audio-recorded | Page 5 |
| Field notes | 20 | Taken during/after interviews | Page 5 |
| Duration | 21 | 45–90 minutes | Page 5 |
| Data saturation | 22 | Discussed/assessed during analysis | Page 6 |
| Transcripts returned | 23 | Offered to participants; none requested | Page 5 |
| **Domain 3: Analysis/findings** | | | |
| Number of data coders | 24 | Two coders (GS and IK) | Page 5 |
| Description of coding tree | 25 | Developed negotiated shared codebook | Page 5 |
| Derivation of themes | 26 | Inductive; participatory thematic analysis | Page 5 |
| Software | 27 | ATLAS.ti | Page 5 |
| Participant checking | 28 | CAB members involved in PTA session | Page 6 |
| Quotations presented | 29 | Presented in Quotes Table and Results | Quotes Table |
| Data/findings consistent | 30 | Consistent across all findings sections | Pages 6-11 |

| | | | |
|---|---|---|---|
| Clarity of major themes | 31 | Clearly presented in Findings | Pages 6-11 |
| Clarity of minor themes | 32 | Discussed in Results and Discussion | Pages 6-11, 11 |

Developed from: Tong A, Sainsbury P, Craig J. Consolidated criteria for reporting qualitative research (COREQ): a 32-item checklist for interviews and focus groups. *International Journal for Quality in Health Care*. 2007. Volume 19, Number 6: pp. 349 – 357

**Once you have completed this checklist, please save a copy and upload it as part of your submission. DO NOT include this checklist as part of the main manuscript document. It must be uploaded as a separate file.**
